# A systematic review on the levels of antibodies in COVID-19 virus exposed but negative newborns: a possible vertical transmission of IgG/ IgM

**DOI:** 10.1101/2020.06.09.20127118

**Authors:** George M. Bwire, Belinda J. Njiro

## Abstract

**Background:** Currently, there is no doubt on human-to-human transmission of Coronavirus Disease 2019 (COVID-19). Now, the debates remain on whether, vertical transmission of Severe Respiratory Syndrome Virus 2 (SARS-CoV-2) and antibodies against the virus do exist. We therefore, conducted a systematic review to determine the immunoglobulin G and M (IgG/IgM) levels among infants born to mothers with COVID-19.

**Methods:** The systematic search was done using PubMed/MEDLINE and Google Scholar database. The research included studies on IgG/ IgM against SARS-CoV-2 among infants born to mother with COVID-19 published in English from December 1, 2019 onwards. Data were extracted by two independent authors in accordance with the Preferred Reporting Items for Systematic Reviews and Meta-Analyses (PRISMA-P) guidelines. We synthesized a narrative from eligible studies and performed two tailed non-parametric Mann-Whitney test to determine and compare the median IgG/IgM levels.

**Results:** In total, 486 abstracts were screened and 63 full-text articles were assessed. Of 63 articles, 6 met the inclusion criteria for qualitative analysis. Two articles were included in quantitative analysis of anti-SARS-CoV-2 IgG/ IgM levels. The median antibody levels was 75.49AU/mL (range: 7.25AU/mL-140.32AU/mL) and for 3.79AU/mL (range: 0.16AU/mL-45.83AU/mL) (P = 0.0041) for anti-SARS-CoV-2 IgG and IgM, respectively.

**Conclusion:** There were high levels of IgG but low IgM against SARS-CoV-2 (using <10 AU/mL as a reference range) among COVID-19 virus exposed but negative newborns. This review suggest a possible natural passive immunity (IgG/ IgM) against COVID-19 virus.

**Impact:** A systematic review of infants born to mothers with COVID-19 was conducted to characterize the magnitude of antibodies generated against SARS-CoV-2 among infants who were vertically exposed to the virus. These findings were necessary to inform the ongoing vaccine development and research on the background natural passive immunity among COVID-19 exposed newborns. Furthermore, evidence are revealing the possibility of vertical transmission of anti-SARS-CoV-2 IgG/ IgM among the exposed newborns who tested negative for the virus.

## Introduction

Since the first case report of Severe Respiratory Syndrome Coronavirus 2 (SARS-CoV-2) in Wuhan, China (1), several studies have reported human-to-human transmission of SARS-CoV-2 leading to Coronavirus Disease 2019 (COVID-19) (2,3). However, evidence on mother to child transmission is still limited, World Health Organization (WHO), reported no evidence on mother-to-child transmission when infection manifests in the third trimester by testing amniotic fluid, cord blood, vaginal discharge, neonatal throat swabs or breastmilk (4). But with the current rising neonatal COVID-19 infections born under well infection control and prevention (IPC) settings, possible vertical transmission is suggested (5–7).

On the other hand, natural passive immunity and detection of specific IgG and IgM antibodies to SARS-CoV-2 in infants born to COVID 19 confirmed mothers have been indicated in some studies where the newborns tested negative for the virus (8). Primarily, IgM and IgG antibodies detection tends to indicate recent exposure/ first line of defense to SARS-CoV-2, whereas the detection of COVID-19 virus IgG antibodies indicates past exposure or recovery from the virus/ disease (9). Moreover, while IgG easily cross the placenta, it is unlikely for IgM to be transferred from mother to fetus due to its large molecular structure (10). However, studies have reported both IgG/ IgM detection in infants born to mothers with COVID-19 but themselves did not contract the disease (11–14). In this regard, a systematic review was conducted to determine the magnitude of IgG/ IgM in infants born to mothers with COVID-19 but tested negative for SARS-CoV-2.

## Methods

### Study design

A systematic review protocol was developed following Preferred Reporting Items for Systematic Reviews and Meta-Analyses Protocols (PRISMA-P) guidelines (15). The protocol was registered in International Prospective Register of Systematic Reviews (https://www.crd.york.ac.uk/prospero/: PROSPERO database registration number: CRD42020188392). This systematic review was conducted to address the following question, “What is the level of anti-SARS-CoV-2 IgG/ IgM among infants born to mothers with COVID-19?”.

### Search strategy

Articles were systematically searched from PubMed/MEDLINE and Google scholar. We also searched from the websites of key healthcare organizations such as WHO and Centre for Disease Control and Prevention. Similarly, a grey literature search (e.g. pre-prints) was done with help of Google. Data from December 31, 2019 to May 18, 2020 conducted in human beings and published in English language. We developed a rigorous systematic search strategy with the help from health sciences librarian who has systematic review experience. The strategy was developed for PubMed/MEDLINE (Additional file 1) using keywords and MeSH (MEDLINE). Keywords such as vertical transmission, passive immunity, antibody, immunoglobulin, mother, child, infant, newborn, SARS-CoV-2 and COVID-19 were used.

### Eligibility criteria and study selection

Two reviewers (GMB and BJN) independently screened the titles and abstracts, and a full-text articles for inclusion. Disagreements on study eligibility were resolved by consensus, and/or a third reviewer was consulted if necessary. The selected studies were included based on pregnant mother with laboratory-confirmed COVID-19 virus infection using quantitative real-time polymerase chain reaction (qRT-PCR) or dual fluorescence PCR, patient pregnant on admission, maternal clinical characteristics including maternal and perinatal outcomes and infection control and prevention measures during and after delivery were assessed before decision to include the article. This review included, letter to the editor, correspondence, editorial, research article (case report, case series, cross-sectional, clinical trial, cohort, case control study) but excluding articles on reviews, opinions and perspectives.

### Data Management

All article citations pooled from database searches were exported into EndNote software version X7 (Thomson Reuters, 2015) for managing the duplicates and for title and abstract screening. Identified publication(s) were analyzed using criteria based on levels of IgG/ IgM against SARS-CoV-2 (Fig. 1).

**Fig. 1.**
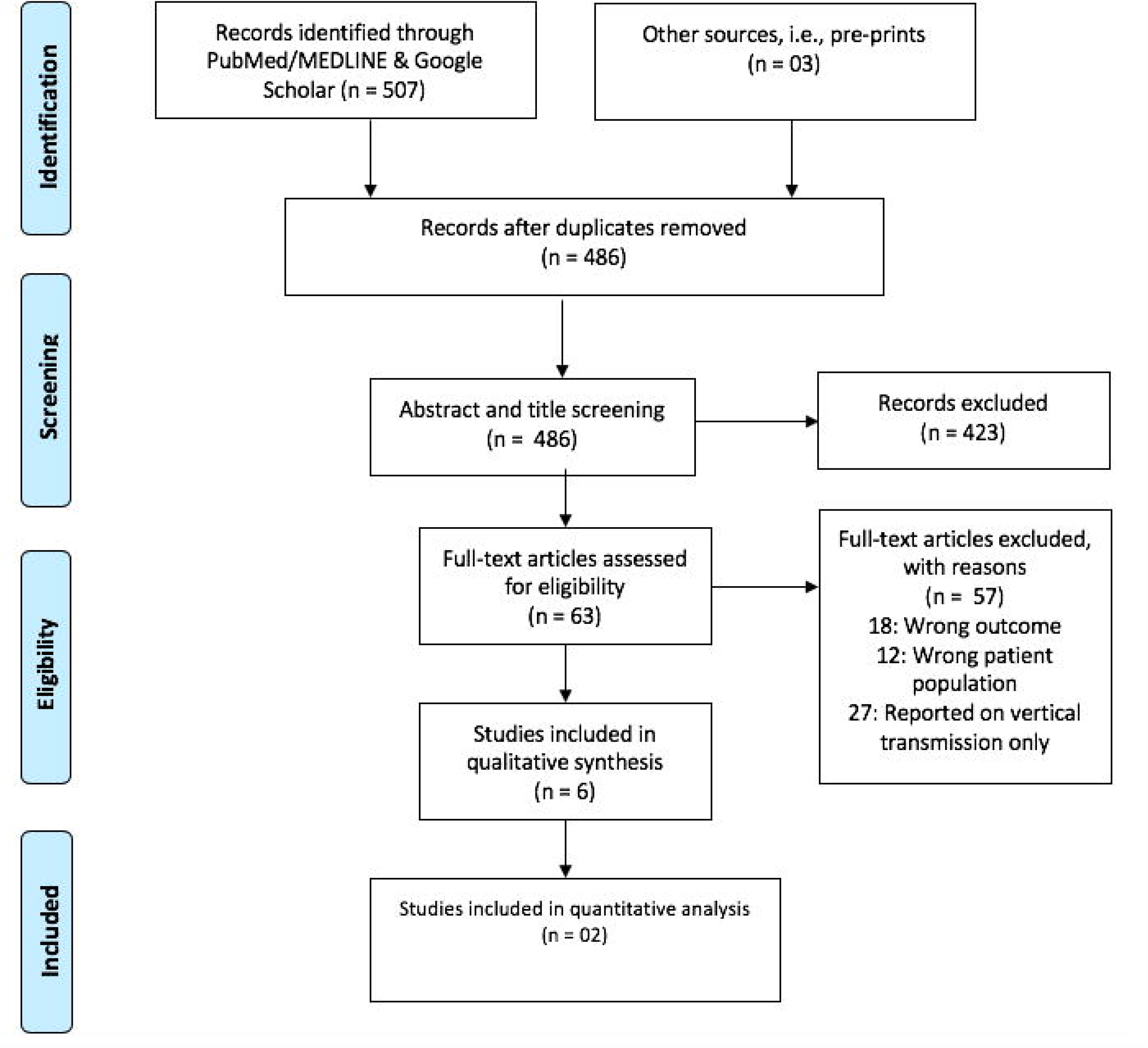
Prism flow chart on how the final studies for inclusion in analysis was obtained

### Data extraction and quality assessment

The reviewers (GMB and BJN) independently extracted the variables of interest from the selected studies. Data extraction was done using Excel spreadsheet 2010 (Microsoft Corporation, Redmond, WA) (Table 1). The primary endpoints were birth outcomes, such as COVID-19 infection, IgM and IgG against SARS-CoV-2. PRISMA-P guideline (15) recommends a quality assessment of the included literature, but most of the extracted studies were case reports with one participant per study and only one retrospective study with 6 participants. In this regard, authors decided not to perform the risk assessment as described previously (16). Data not enough to perform heterogeneity and meta-analysis. However, reviewers were stringent on inclusion criteria, to minimize the methodological errors.

**Table 1:** Characteristics of studies included for analysis

| Author | Country | Study design | Mode of delivery | Gestation age at delivery (weeks) | COVID-19 virus newborn status | Antibody levels for SARS-CoV-2 in newborns |  | Method for Ig detection/quantification |
| --- | --- | --- | --- | --- | --- | --- | --- | --- |
|  |  |  |  |  |  | IgG (AU/mL) | IgM (AU/mL) |  |
| Alzamora et al.,2020 (19) | Peru | Case report | Cesarean | 33 | Positive | Not detected | Not detected | Not reported |
| Buonsenso et al.,2020 (11) | Italy | Case report | Cesarean | 38<sup>+3</sup> | Negative | Weakly positive but not quantified | Not detected | Not reported |
| De Socio et al.,2020 (12) | Italy | Case report | Vaginal | 40 | Negative | Strongly positive but not quantified | Weakly detected but not quantified | COVID-19 IgG/IgM fast assay (Screen Italia Srl, Perugia, Italy) |
| Zeng et al.,2020 (17) | China | Retrospective study of 6 infants | Cesarean | Not reported | Negative | 125.5 | 39.6 | SARS-CoV-2 IgG and IgM (CLIA assays Kit, YHLO) |
|  |  |  |  |  |  | 113.91 | 16.25 |  |
|  |  |  |  |  |  | 75.49 | 3.79 |  |
|  |  |  |  |  |  | 73.19 | 1.9 |  |
|  |  |  |  |  |  | 51.38 | 0.96 |  |
|  |  |  |  |  |  | 7.25 | 0.16 |  |
| Xiong et al.,2020 (18) | China | Case report | Vaginal | 38<sup>+4</sup> | Negative | Not detected | Not detected | IgG and IgM (SARS-CoV-2;INNOVITA, China). |
| Dong et al.,2020 (14) | China | Case report | Cesarean | 34<sup>+2</sup> | Negative | 140.32 | 45.83 | Not reported |

### Summary measures and synthesis of results

Since the extracted data on IgG/ IgM levels were not normally distributed. We performed two tailed non-parametric Mann-Whitney test to determine and compared the median antibody levels (Prism 7 software; GraphPad Software, USA). The narrative was written by the lead reviewer (GMB) and then checked independently by reviewer (BJN). The variables that were missing from included articles were recorded as not reported. No statistical test was performed for missing data.

## Results

### Characteristics of the analyzed studies

A total of 510 literatures were pooled after a systematic search. Out of 510 articles, 63 (12.4%) were eligible for full-text assessment. Of 63 full-text assessed articles, 6 (9.5%) reported about IgG/IgM against SARS-CoV-2 where 2 articles had a quantified (reported the metric values) IgG/IgM levels (14,17), 2 articles reported unquantified detection of IgG/IgM (11,12). Two articles reported that no IgG/ IgM was detected from the newborns samples tested (18,19). Except for one study which was a retrospective study (17), the remained studies (11,12,14,18,19) included for qualitative analysis were case reports where one study reported an infant who tested positive for COVID-19 virus (19). For the rest of the articles infants tested negative for the virus. In terms of mode of delivery, 3 delivered through vaginal route and 3 by cesarean route. Preterm deliveries were reported in two studies (14,19). Three studies reported an assay employed in IgG/ IgM quantification (12,17,18).

### IgG and IgM levels against SARS-CoV-2

The median antibody levels detected in COVID-19 exposed newborns who tested negative for the virus after delivery but were born to mothers with COVID-19 were 75.49AU/mL (range: 7.25AU/mL-140.32AU/mL) and for 3.79AU/mL (range: 0.16AU/mL-45.83AU/mL) (P = 0.0041) for anti-SARS-CoV-2 IgG and IgM, respectively. Study by Alzamora et al., (19) found that, newborns who tested positive for SARS-CoV-2 had not antibodies against the virus during the time of sample collection. Presence of antibodies was not detected in an infant born at 34^+2^ weeks by vaginal route (18). This child tested negative for COVID-19 virus too. There was strongly positive IgG and weakly IgG in Italian baby delivered by cesarean route (11) while Buonsenso et al., (11) reported weakly positive IgG and absence of IgM.

## Discussion

This review was conducted to determine the magnitude of IgG/ IgM against SARS-CoV-2 in infants born to mothers with COVID-19. Overall, six articles reported on antibodies detection for SARS-CoV-2 in newborns, five of them infants tested negative for the virus (11,12,14,17,18) whilst one study a child tested positive for the virus (19). Using the reference range of <10 AU/mL (17), this study found high levels of antibodies (75.49AU/mL) for IgG and low levels (3.79AU/mL) of IgM against SARS-CoV-2 among the exposed but negative newborns.

High levels of IgG were similar from other findings which described its small molecular structure (10) and ability to start crossing the placenta at the beginning of the end of the second trimester (17). Generally, IgM are highly expressed as the first line antibodies during the time where the disease is active whereas IgG detection usually indicates the long time infection or recovery from the past infection (9). Furthermore, IgM are large in molecular structure making them unlikely to cross the placenta (10). However, detection of IgM in the recent studies (12,14,17) suggest the possibility that, anti-SARS-CoV-2 IgM can cross the placenta.

Immunological experience form other respiratory infection such as influenza, indicated that natural transplacental influenza antibodies protect infants during the first few months of life (20). Additionally, artificial maternal influenza antibodies significantly reduced the rate of laboratory-confirmed influenza in the infants (21). That experience can be employed to study the generated IgG/ IgM against SARS-CoV-2 and find out their protective effect to the newborns. In this review one case reported absence of IgG/IgM against the SARS-CoV-2 but infant tested positive for the virus (19). Also, study reported negative serological test from his mother on the date of birth. The reason for this can be the explained from the recently acquired infection where IgM (first antibodies to be produced) seroconverts after day 5 from symptom onset (22).

Given the time since the first case of COVID-19 (December 31, 2019), this study is limited by the small number of participants obtained from few pooled studies. However, in the fight against the limited known COVID-19 pandemic, every available scientific information should be timely employed to guide the ongoing therapeutic and vaccine research and development.

## Conclusion

There were high levels of IgG but low IgM against SARS-CoV-2 (using <10 AU/mL as a reference range) among COVID-19 virus exposed but negative newborns. This review suggest a possible vertical transmission of anti-SARS-CoV-2 IgM. These findings were necessary to inform the ongoing vaccine development and research on the background natural passive immunity among COVID-19 exposed newborns. In addition, evidence are revealing the possibility of vertical transmission of anti-SARS-CoV-2 IgG/ IgM among the exposed newborns but who tested negative for the virus.

## Data Availability

Data used to draw this conclusion are available from the corresponding author without restriction

## Acknowledgment

We thank Deodatus Sabas, Librarian at Muhimbili University of Health and Allied Sciences, Dar es Salaam, Tanzania for his assistance and support in refining the search strategy. Authors acknowledge the training support received from Muhimbili University of Health and Allied Sciences through Systematic Review and Meta-analysis workshop funded by Swedish International Development Cooperation Agency, Sweden.

## Authors contributions

Conceptualization, Data curation, Formal analysis, Methodology, Manuscript drafting: George M. Bwire.

Data curation, Formal analysis, Methodology, Manuscript review and editing: Belinda J. Njiro. Both authors have read and approved the final version of manuscript.

## Data availability

All datasets used and analyzed in this study are provided in the reference list and its supplementary material (Additional file 1 & 2).

## Ethics statement

Not applicable

## Table and Figure Legends

**Additional file 1:** PubMed/ MEDLINE search strategy and results

**Additional file 2**: Prisma populated checklist

